# Performance of low-cost non-invasive blood markers of liver cirrhosis in adults with chronic hepatitis B infection with and without comorbid alcohol use in Zambia

**DOI:** 10.1101/2024.04.23.24306219

**Authors:** Sydney Mpisa, Morris Kahere, Annie Kanunga, Michael Vinikoor, Edford Sinkala

## Abstract

**Background:** Diagnosis of liver cirrhosis in patients with chronic hepatitis B is challenging given rare use of biopsy. In low and middle-income countries, transient elastography (TE), a recommended non-invasive imaging test for cirrhosis is rarely accessible. We therefore investigated the performance of multiple low-cost and more accessible blood-based liver fibrosis markers in patients with chronic hepatitis B infection in Zambia. As alcohol use complicates the assessment and outcomes of hepatitis B, we also considered alcohol use patterns in our evaluation.

**Methods:** We performed a hospital-based cross-sectional study, in Lusaka, Zambia, among consecutive treatment-naive adults with chronic hepatitis B mono-infection (i.e., HIV-negative) presenting to our hospital. The reference test for cirrhosis was TE of >/=9.6 kPa. Low-cost markers were the AST-to-platelet ratio index (APRI) at recommended threshold >2, as well as lower proposed alternative thresholds for Africa, >0.5 and >0.65, AST/ALT ratio and FIB-4 index >3.25. We evaluated the performance of each marker versus TE. In a secondary analysis, we evaluated marker performance in participants with current alcohol use versus lifetime abstinence.

**Results:** Among 239 adults with HBV mono-infection analyzed, the mean age was 34.7 years and 53 (22.2%) reported current alcohol use. The prevalence of cirrhosis by TE was 16.3% (95% CI: 11.87-21.63). The area under the receiver operating characteristic curve was 0.83, 0.80, 0.79 and 0.73 for FIB-4, APRI >0.5, APRI >0.65 and APRI >2 respectively. Virtually all indices performed less well in people with current alcohol use.

**Conclusion:** These data support the adoption of a lower APRI threshold in Africa, and the use of the FIB-4 index, for diagnosis of cirrhosis among patients with chronic hepatitis B infection. The currently-recommended APRI threshold may exclude people with cirrhosis who need antiviral therapy. Clinicians adopting these markers should screen for alcohol use and consider re-assessment of cirrhosis after alcohol reduction.

## Introduction

According to WHO 2023 report approximately 296 million individuals worldwide were affected by hepatitis B virus in 2019 (WHO, 2023). In sub-Saharan Africa, where hepatitis B is endemic, the overall sero-prevalence of hepatitis B surface antigen remains as high as 6.1% (Spearman et al., 2023). In Zambia, the sero-prevalence of HBV was reported as 5.6% in 2016 (Franklin et al., 2018). The substantial prevalence of hepatitis B in sub-Saharan Africa carries clinical ramifications, notably liver cirrhosis, and hepatocellular carcinoma (HCC). Mozambique stands out as one with the highest HCC incidences, primarily linked to HBV (Petruzziello, 2018).

Once a person is diagnosed with chronic HBV infection, indications for antiviral therapy are considered. A major factor in treatment decisions is the presence or absence of liver cirrhosis; however, diagnosing cirrhosis can be challenging since the ‘gold standard’ is a liver biopsy, which is invasive and is associated with sampling error (Regev et al., 2002). In 2015, based mostly on data from Asia and hepatitis C virus infections, WHO recommended non-invasive markers of fibrosis, transient elastography, an imaging tool, and AST-to-platelet ratio index (APRI) score >2(Organization, 2015). While transient elastography (TE) is validated for diagnosing liver fibrosis (Kim et al., 2009), its limited availability in most hospitals underscores the need for affordable and accessible methods for liver fibrosis assessment in low-resource settings. Because of the low cost and ability to be performed with existing equipment (unlike elastography), the APRI became widely adopted in low and middle-income countries for HBV management; however, subsequent studies in Africa revealed that this cut-off missed some patients requiring treatment (Aberra et al., 2019). Achieving WHO’s 2030 goals of eliminating HBV demands decentralizing and simplifying treatment for primary care physicians and this calls for simpler ways of assessing liver fibrosis. More recently APRI >0.65 was proposed based on analysis of African data (Johannessen et al., 2022).

In this study, we sought to evaluate the performance of low-cost non-invasive liver fibrosis markers (APRI, at current and proposed thresholds, FIB-4 index and other similar indices) in predicting liver fibrosis among patients with chronic HBV infection in sub-Saharan Africa. First, we estimated the prevalence of liver cirrhosis among patients with chronic HBV mono-infection who are treatment naïve. We then assessed the degree to which the non-invasive liver fibrosis markers perform in diagnosing liver cirrhosis using transient elastography as the acceptable ‘gold standard’ among patients with chronic HBV mono-infection. We also sought to identify demographic, social and clinical predictors of cirrhosis. Because aspartate aminotransferase (AST) and alanine aminotransferase (ALT) are non-specific measures that can be increased with alcohol use, which is common in Zambia and the region, we also stratified analyses by alcohol use patterns.

## Methods

### Study design, setting and participants

This study, conducted from 15 April 2022 to 24 January 2023 at the University Teaching Hospital (UTH) in Lusaka, Zambia, was an analytical cross-sectional investigation. UTH is a tertiary hospital in Lusaka, Zambia. The research involved reviewing medical records of adult male and female patients diagnosed with chronic hepatitis B infection who provided consent and were attendees of our hepatitis clinic. The hepatitis clinic at UTH is done weekly, staffed by trainees and consultant hepatologists and infectious disease specialists. Overall, nearly 500 persons with HBV are being followed up at our hepatitis clinic. UTH is a referral centre and has enhanced access to HBV expert doctors and diagnostics, it has a large catchment population, including most of Lusaka. This study was conducted in accordance with the Declaration of Helsinki and was reported in compliance with the Strengthening the Reporting of Observational studies in Epidemiology (STROBE) checklist and explanation.

### Participants and eligibility criteria

Participants included in this study were adults aged 18 years and above and had a positive result for Hepatitis B viral infection, defined by the presence of hepatitis B surface antigen (HBsAg) in blood. We excluded individuals who tested positive for HIV, those unwilling or unable to provide consent, those displaying signs of possible acute hepatitis (elevated ALT levels exceeding three times the upper limit of normal or jaundice), those with suspected hepatocellular carcinoma, individuals currently on anti-tuberculosis drugs at the time of recruitment (that may alter liver transaminases), and those who had taken antiviral drugs for HBV infection for more than a month. These criteria were applied to ensure a targeted study population focused on chronic Hepatitis B, while excluding individuals with potential confounding factors or acute conditions that could impact study outcomes.

### Sampling and sample size estimation

Consecutive enrolment of all consenting participants with hepatitis B viral infection who were treatment naïve was adopted as it was deemed appropriate. Due to the cross-sectional nature of the study, the sample size was determined based on prevalence calculation. The minimum sample size was computed using the prevalence formula, which was estimated based on the following parameters: the required level of confidence, set at 95% (corresponding to a standard normal distribution value of 1.96); an assumed prevalence for liver fibrosis in the study setting, using 20% or 0.2 from a previous study done in Zambia(Vinikoor et al., 2018); and a desired degree of accuracy, specified at 5% or 0.05, representing the precision sought in the width of each arm of the confidence interval. The required sample size for the study was 246.

### Non-invasive serum markers of fibrosis

The use of serum markers of fibrosis has been an area of interest because liver cirrhosis requires histological confirmation, and this comes with its own challenges. APRI score is a ratio of the patient aminotransferase and platelet which is calculated as follows: **[(AST/upper limit of the normal AST range) X 100]/Platelet Count**. The calculated score can be used to diagnose liver cirrhosis with different cut-off scores being suggested in literature. WHO guidelines in 2015 adopted an APRI score >2 to diagnose liver cirrhosis. FIB-4 index (Fibrosis 4 index) is a test derived from the Apricot database, the FIB-4 index combines biochemical values (platelet count, ALT and AST) and age as follows:

**FIB-4 = Age (years)×AST (U/L)/ [PLT (10**^**9**^**/L) ×ALT**^**1/2**^ **(U/L)]**. A threshold value of 3.25 has a positive predictive value for the diagnosis of extended fibrosis of 65% (Vallet-Pichard et al., 2007). Of note, the FIB-4 does not have a specific threshold for cirrhosis, but we opted to evaluate it as it is low cost and has potential for implementation in Africa. AST/ALT ratio is commonly used in alcoholic liver disease where a ratio >2 is predictive but it has also been used to assess the extent of fibrosis in viral hepatitis. A ratio of AST/ALT above 1 has been used to diagnose liver cirrhosis though its performance has been found to be unreliable.

### Study instruments and data collection

Data were collected from the patients and patient’s medical records by a trained and experienced research assistant (a nurse), using a tool that was developed and pilot tested by the principal investigator prior to its final use. This tool collected information on participant’s demographic characteristics, medical history, alcohol history and laboratory investigations, including HBV DNA, HBeAg, and tests used in the low-cost fibrosis scores. The data collected within 30 days of the transient elastography were obtained for valid comparison of the laboratory results and the TE values. The principal investigator checked/verified all the collected data to minimize bias and recording errors and to ensure accuracy and reliability of the data collection process.

### Ethical consideration

Gatekeeper permission was sought from the participating institutions prior to seeking signed informed consent from the individual participants. The study was submitted to the University of Zambia Biomedical Research Ethics Committee and National Health Research Authority for ethical approval. As it was an observational study, no interventions were instituted, patients continued to receive their usual care with no extra costs to the participants. Participation of all respondents in the study were all strictly voluntary. Respect, dignity and freedom of each individual participating in the study was granted. To guarantee the anonymity of each participant, the names of respondents, their addresses or other identifying information were not included in the data collection. All participants were required to provide written informed consent, those who couldn’t consent for any reason were excluded from the study. The data was kept secure by the investigator and a coding system used to maintain confidentiality.

### Statistical analysis

The data cleaning and analysis process utilized IBM Statistical Package for Social Sciences (SPSS) software version 27.0 for Windows (IBM Corp., Armonk, NY, USA). Descriptive statistics were employed, presenting frequencies and percentages for categorical variables, mean and standard deviation for age, and median with interquartile range for liver cirrhosis diagnostic markers. Alcohol use was categorized as never (i.e., lifetime abstinence), current use, or past use, because abstainers with past vs. no lifetime alcohol use may have different degree of liver disease. Sensitivity and specificity analyses were conducted using crosstabs to assess the performance of low-cost markers. The diagnostic performance of low-cost markers was expressed in terms of the sensitivity (se), specificity (sp) and area under the ROC curve (AUROC). Optimised cut-offs for the low-cost markers and for liver cirrhosis were obtained analysing the AUROC at the maximum of total sensitivity and specificity. Dichotomous (1 for the presence of liver cirrhosis and 2 for NO cirrhosis) low-cost markers considered in the analysis were APRI >0.65, APRI >0.5, APRI >2, FIB-4 index >3.25, AST/ALT>1, AST/ALT>2 and Platelet count<150. The TE score categories were defined as 0 (absence of cirrhosis, less than 9.6 kPa) and 1 (presence of cirrhosis, 9.6 kPa or greater) (Lemoine et al., 2016).

## Results

A total number of 285 patients were screened for inclusion in the analysis; 46 were excluded based on being on antiviral therapy for more than a month, non-availability of TE results, and/or suspected acute HBV infection (Figure 1), yielding an 83.9% eligibility rate. Male to female ratio was 3:2 with a mean age (±SD) of 34.7 (± 10.6) years. A little less than half of the participants had never drank alcohol in their life (46.5%) by report, with 23.0% and 30.4% being current and former drinkers, respectively. Variables with missing values are shown in table 1.

**Table 1.** General characteristics of the participants analyzed (N = 239)

| Variable |  | Value | Missing data |
| --- | --- | --- | --- |
| Age (years) mean ± SD | | 34.7 (10.6) | |
| Sex, N (%) |  |  | No missing values |
|  | *Female* | 91 (38.1) |  |
|  | *Male* | 148 (61.9) |  |
| Alcohol consumption pattern, N (%) |  |  | 9 |
|  | *Current alcohol use* | 53 (23.0) |  |
|  | *Former alcohol use* | 70 (30.4) |  |
|  | *Lifetime abstainer* | 107 (46.5) |  |
| ALT (U/L) median (IQR) |  | 25.0 (19-41) | 6 |
| AST (U/L) median (IQR) |  | 28.5 (24-40.8) | 3 |
| Platelet count (/μL) median (IQR) | | 213.0 (164-260) | No missing values |
| Transient Elastography score (kPa) median (IQR) |  | 5.6 (4.5-7.4) | No missing values |
| HBV DNA results (%) |  | 207 (86.6) | 32 (13.4) |
|  | HBV DNA VL<2000IU/ml | 124 (59.9) |  |
|  | HBV DNA VL>2000IU/ml | 83 (40.1) |  |
| HB 'e' Antigen results (%) |  | 101 (42.3) | 138 (57.7) |
|  | HB 'e' antigen positive (%) | 21 (20.8) |  |
|  | HB 'e' antigen negative (%) | 80 (79.2) |  |
| APRI median (IQR) |  | 0.4 (0.3-0.7) | 3 |
| AST/ALT ratio median (IQR) |  | 1.2 (0.9-1.5) | 8 |
| FIB-4 median (IQR) |  | 0.9 (0.6-1.5) | 8 |

*SD* standard deviation *IQR* interquartile range, *N* number, *ALT* alanine aminotransferase, *AST* aspartate aminotransferase, *APRI* AST to platelet ration index, *Fib-4* Fibrosis 4

**Figure 1:**
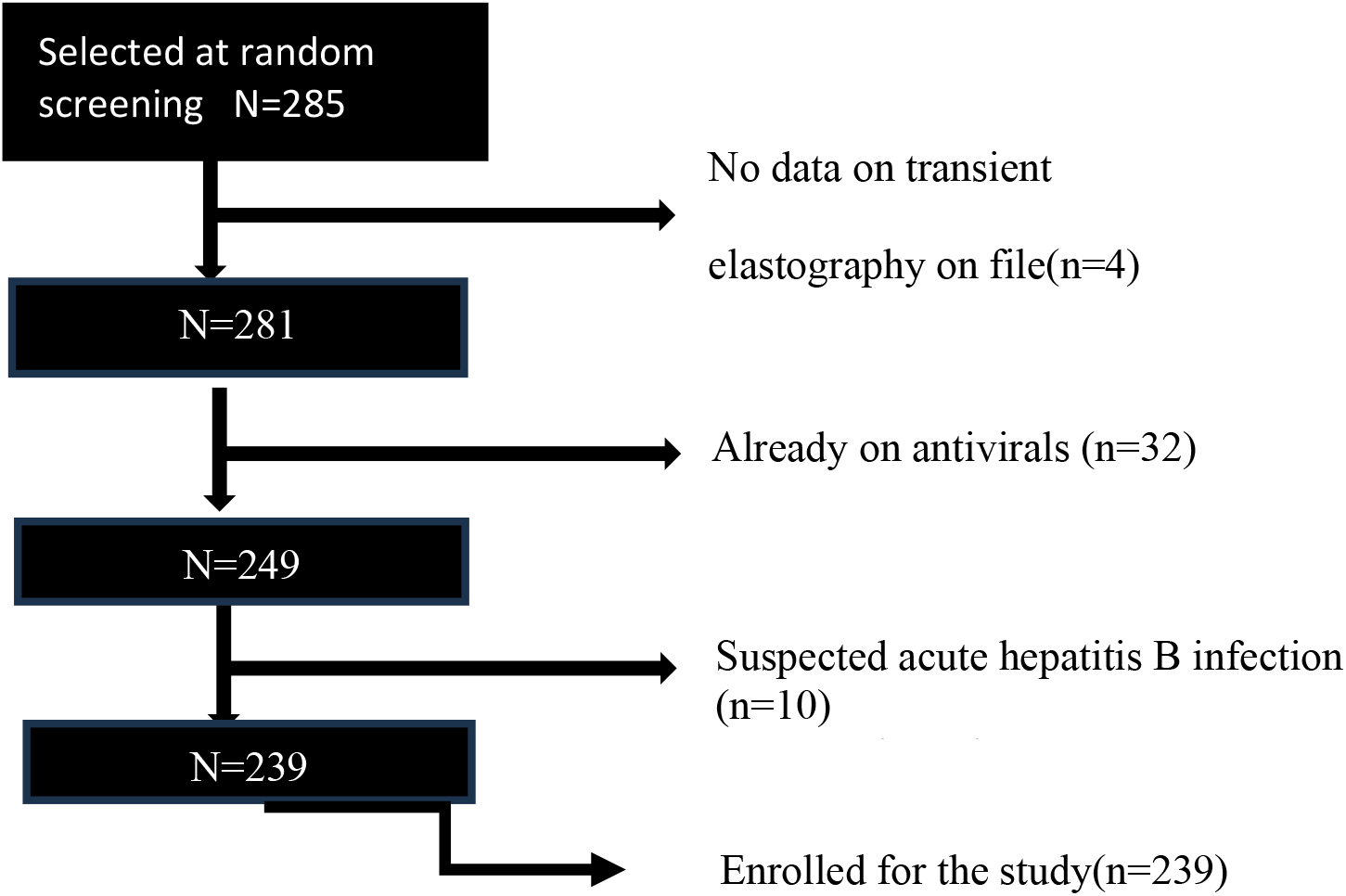
study recruitment

Based on TE cut-off of 9.6kPa, (Lemoine et al., 2016) the prevalence of liver cirrhosis in the analysis group was 16.3% (95% CI: 11.9 – 21.6). Among the 239 participants 101 had hepatitis e antigen results with 21 (20.8%) of those having positive results. A total of 207 participants had hepatitis B DNA viral load results with 83 (40.1%) having a value more than 2,000 IU/ml. We conducted a bivariate analysis (chi-square test) and it showed that APRI >0.65, APRI >0.5, APRI >2, FIB-4 index >3.25, platelet<150 and AST/ALT>2 where significantly associated with cirrhosis except for the AST/ALT cut-off >1, *x*^2^=0.12, df=1, p=0.73. Among all the APRI thresholds, APRI cut-off 2 showed the greatest association (x^2^=97.31, df=1, p<0.001) with cirrhosis, and the association had a strong effect size (Cramer’s V=0.64). FIB-4 index also had strong association with cirrhosis (x^2^=88.73, df=2, p<0.001) with a strong effect size (Cramer’s V=0.62).

### Performance of low-cost markers to diagnose cirrhosis

APRI >0.65 had good sensitivity and specificity with values of 76.5% and 84.8% respectively whilst AST/ALT ratio>2 had the least sensitivity of 20% showing a false negativity of 80% (Table 2). The current WHO recommended APRI cut-off 2 had good specificity (99%) but has very high false negativity 48.7% which means a number of patients can be missed thus miss out on treatment. FIB-4 index had the least false positivity at 3.1% and false negativity 20.5% and when plotted on the area under curve it had the greatest AUROC as shown in Figure 2.

**Table 2.** Performance of low-cost markers in diagnosis of liver cirrhosis.

| Test performance | FIB-4 >3.25 | APRI>0.5 | APRI>0.65 | APRI>2 | AST/ALT>1 | AST/ALT>2 | Platelet<150 |
| --- | --- | --- | --- | --- | --- | --- | --- |
| Sensitivity (%) | 51.3 | 79.5 | 76.9 | 51.3 | 40.9 | 20 | 66.7 |
| Specificity (%) | 83.3 | 80.7 | 84.8 | 99 | 62.2 | 94.8 | 89.0 |
| False +ve (%) | 3.1 | 19.3 | 15.2 | 1 | 59.1 | 5.2 | 11.0 |
| False -ve (%) | 20.5 | 20.5 | 23.1 | 48.7 | 37.8 | 80 | 33.3 |

*FIB-4*, *APRI* AST platelet ratio index, *AST* aspartate aminotransferase, *ALT* alanine aminotransferase.

**Figure 2:**
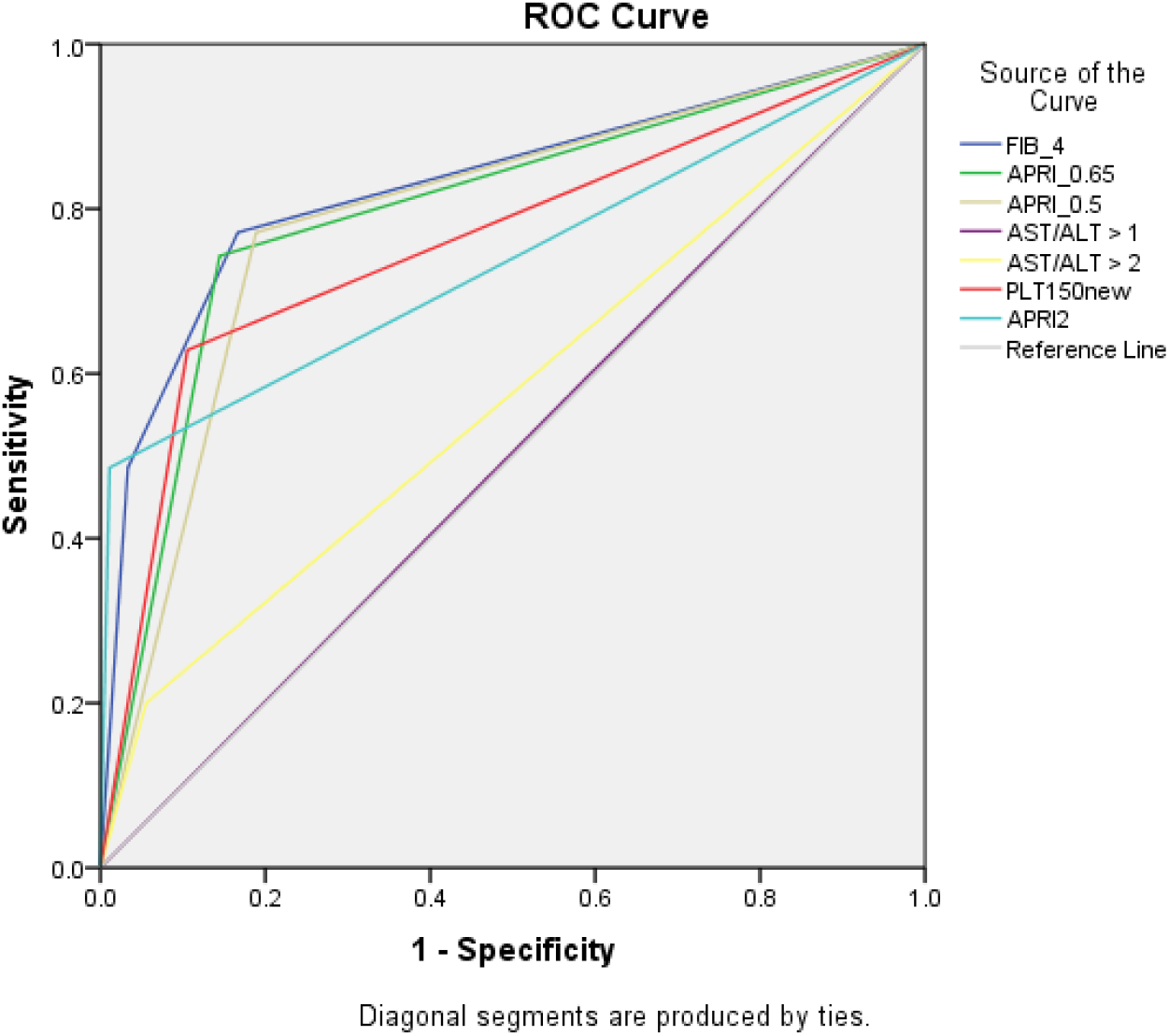
ROC curve showing performance of different markers in predicting cirrhosis based on transient elastography.

### Receiver-Operating Characteristic Analysis

Figure 2 and table 3 delineates outcomes from a receiver operating characteristics curve (ROC) and an area under the curve (AUC) analysis assessing the diagnostic efficacy of APRI>0.65, APRI>0.5, APRI>2, Platelet <150, AST/ALT > 1, FIB-4 index>3.25, and AST/ALT > 2. AUC, a metric gauging diagnostic test performance, reveals values proximal to 1 signify superior efficacy. Results indicate that FIB-4 index exhibited the highest performance (AUC = 0.83, 95% CI: 0.74–0.91), surpassing APRI >0.5 (AUC = 0.80, 95% CI: 0.72–0.88) and APRI >0.65 (AUC = 0.78, 95% CI: 0.69–0.87). Conversely, AST/ALT ratios, both > 1 (AUC = 0.53, 95% CI: 0.44–0.63) and > 2 (AUC = 0.53, 95% CI: 0.42–0.63), demonstrated relatively inferior performance. In summary, FIB-4 >3.25 emerges as the optimal performer, while APRI >0.5 cut-off exhibits marginally better efficacy than APRI >0.65.

**Table 3:** Area under the ROC curve (AUROC)

| Test result variables | AUROC | Standard error | P value | Lower bound 95% CI | Upper bound 95% CI |
| --- | --- | --- | --- | --- | --- |
| APRI >0.65 | 0.78 | 0.05 | 0.000 | 0.69 | 0.87 |
| APRI >0.5 | 0.80 | 0.04 | 0.000 | 0.72 | 0.88 |
| AST/ALT>1 | 0.53 | 0.05 | 0.507 | 0.44 | 0.63 |
| FIB-4 >3.25 | **0.83** | 0.04 | 0.000 | 0.74 | 0.91 |
| AST/ALT>2 | 0.53 | 0.05 | 0.628 | 0.42 | 0.63 |
| Platelet <150 | 0.76 | 0.05 | 0.000 | 0.66 | 0.86 |
| APRI >2.0 | 0.73 | 0.06 | 0.000 | 0.63 | 0.85 |

As alcohol both causes cirrhosis and can raise the AST and ALT, we performed a stratified analysis to determine if alcohol consumption pattern affected the diagnostic performance of the low-cost cirrhosis markers. This subgroup analysis constituted at total sample of 230 participants, comparing 53 people with current alcohol user and 107 with lifetime abstinence. As seen in Table 4 diagnostic performance differed with alcohol use. The area under curve for FIB-4 index was reduced in current alcohol drinkers at (0.70) compared to (0.80) in participants who never took alcohol. The trend was similar in all the other tests (APRI >0.5, APRI >0.65, Platelet<150) but APRI >2 showed opposite results with improved area under curve in current alcohol drinkers (0.82) compared to (0.76).

**Table 4:** Effect of alcohol on the performance of the non-invasive diagnostic tests.

| Diagnostic test | Alcohol status | Sensitivity % | Specificity % | AUROC | 95% CI |
| --- | --- | --- | --- | --- | --- |
| APRI>0.65 | Current | 66.7 | 82 | 0.74 | 0.42-1.00 |
|  | Never | 90.6 | 75 | 0.80 | 0.67-0.94 |
| APRI>0.5 | Current | 66.7 | 76.0 | 0.70 | 0.38-1.00 |
|  | Never | 75.0 | 85.9 | 0.78 | 0.64-0.91 |
| APRI>2 | Current | 66.7 | 98 | 0.82 | 0.49-1.00 |
|  | Never | 50.0 | 100 | 0.74 | 0.58-0.90 |
| FIB-4 >3.25 | Current | 0.0 | 77.6 | 0.70 | 0.38-1.00 |
|  | Never | 55.0 | 86.9 | 0.80 | 0.67-0.95 |
| Platelet <150 | Current | 33.3 | 92.0 | 0.61 | 0.24-0.97 |
|  | Never | 55.0 | 94.3 | 0.74 | 0.59-0.89 |

## Discussion

At a specialized HBV clinic in Lusaka, Zambia, in a consecutive sample of treatment naive patients with chronic HBV, 16.3% had cirrhosis based on TE. The most widely used (WHO recommended) low-cost index, APRI >2, performed less well (AUROC:0.73) compared to other APRI thresholds (APRI >0.5 and APRI >0.65, with AUROC of 0.80 and 0.78), respectively. FIB-4 had the highest AUROC (0.83) for cirrhosis, using a threshold of >3.25 which was previously proposed for fibrosis (i.e., pre-cirrhosis) in people with HCV in the United States guidelines (Sterling et al., 2006). Finally, in a secondary analysis, we observed that across nearly all blood indices, accuracy was reduced in those individuals with comorbid alcohol use. This project supports the implementation of HBV care in Africa and underscores the value of using local data to inform practice guidelines.

The age of 40 years and above seems to be a risk factor for cirrhosis in patients with chronic HBV according to a study done in France (CADRANEL et al., 2007). Our study revealed the median age of 35, therefore there is need to be vigilant in detecting cirrhosis early in our patients. Our study had more males almost twice as many as females infected with chronic HBV, similar to the findings in South Africa (Kew, 2008). The sex disparity of HBV-related liver diseases has been noticed for a long time, which could be attributed to sex hormone effects, other than gender behaviours or environmental impact (Wang et al., 2015). Previous large cohort and 12-year follow-up studies showed that biological factors are more important than environmental or behavioural factors. The higher HBV in men indicates that females develop HBV antibodies faster than males, which helps them to clear HBsAg early. Additionally, the female sex hormone, oestrogen, may be a protective factor for females and androgen as a risk factor for men contributing to higher HBV infections (Shimizu et al., 2007, Sun et al., 2017).

In our evaluation of a range of low-cost markers to diagnose cirrhosis, to circumvent the need for elastography and biopsy, we found in Zambia that APRI >0.5, APRI >0.65, FIB-4 and low platelet <150 had ability to diagnose cirrhosis. Notably, APRI score >2, based on WHO guidelines was widely adopted including in Zambia, but performed poorly in our study and two others in Ethiopia (Dusheiko and Lemoine, 2019, Aberra et al., 2019). This is clinically important as it could lead to missed opportunities to start treatment (i.e. too many false negatives) and this finding was also similar in our study where we found APRI >2 with 48.7% false negativity, compared to 20.5% and 23.1% for APRI >0.5 and APRI >0.65 respectively. Our results agree with the latest findings from most studies which show that lower thresholds like APRI >0.65 has better performance in predicting liver cirrhosis (Jain et al., 2015). A recent meta-analysis looked at the performance of APRI thresholds among patients with chronic hepatitis B virus infection in sub-Saharan Africa also found that APRI>0.65 had better performance in predicting cirrhosis compared to the WHO recommended APRI>2(Johannessen et al., 2022). In our study APRI >0.5 had slightly better area under the curve than APRI >0.65 in diagnosis of cirrhosis based on TE cut-off 9.6kPa with areas of 0.80 and 0.78 respectively. Our study showed that FIB-4 index had the greatest area under curve at 0.83 which is closest to 1 compared to the other non-invasive tests assessed in our study. This FIB-4 threshold >3.25 was proposed for fibrosis (i.e., pre-cirrhosis), but our data suggest it may have a role for cirrhosis detection too. Our results showed that a platelet cut-off 150 was a good predictor of liver cirrhosis based on TE results. The Baveno VI consensus has adopted a platelet cut-off 150 as a predictor for clinically significant portal hypertension (Moctezuma-Velázquez and Abraldes, 2017). Based on the results from this study we may adopt a platelet cut-off 150 as another low-cost predictor of cirrhosis. The adoption of isolated low platelet count as a predictor of liver cirrhosis can be significant in remote areas where doing liver enzymes may be a challenge. The use of platelet count as a predictor of cirrhosis in chronic HBV infection should be applied in clinical context since some regions in Zambia are endemic to schistosomiasis, which causes hypersplenism thus causing a low platelet count in the absence of cirrhosis. Other common causes of a low platelet include such as malaria, bacterial and viral infections should be considered.

Finally, we observed that across nearly all measures, current alcohol intake reduced accuracy of the tests. Of the 239 enrolled participants about half had history of alcohol use either current or past history, as such the effect of alcohol on the non-invasive markers of fibrosis was an important research question. APRI thresholds >0.5 and >0.65 had reduced areas under curve in those with current alcohol history with values dropping from 0.80 to 0.74 for APRI >0.65 and 0.78 to 0.70 for APRI >0.5 when compared to lifetime abstainers. The trend was similar even for FIB-4 index >3.25 which had an area of 0.70 in current alcohol drinkers compared to 0.81 in those who had never taken alcohol. This is an important finding as Africa has seen a rise in unhealthy alcohol use, and these data show that alcohol overlaps with the HBV epidemic in Southern Africa. We know that alcohol interferes with platelet production and therefore this could explain the poor performance of non-invasive markers in predicting cirrhosis (Ballard, 1997). Alcohol also causes oxidative liver injury resulting in elevation the aminotransferases and it increases the progression to liver cirrhosis among patients with chronic hepatitis B infection(McMahon, 2009). A study done in 2006 showed APRI had low sensitivity and specificity for the diagnosis of significant fibrosis in patients with alcoholic liver disease, including patients who had hepatitis C (Lieber et al., 2006). On the basis of our data, guidelines should strongly advocate that people with HBV be screened for alcohol use and that clinicians may consider re-assessing for low-cost biomarkers of cirrhosis once alcohol use has reduced.

### Limitation of this study

While the study had important strengths including a moderate sample size of adults with chronic HBV mono-infection, use of transient elastography, and good clinical characterization including related to alcohol use comorbidity and HBV DNA levels, it also had limitations. The study was a single site study done at a referral centre, which is enriched of people with cirrhosis. Thus, the prevalence of cirrhosis in this sample is likely be higher than in the general population. Also, alcohol was not objectively quantified or confirmed with a biomarker, which might bring some bias. For that reason, we excluded people with past use, whom we suspected may be falsely reporting abstinence, and also overcoming the recall bias.

## Conclusion

While low-cost liver fibrosis markers should be encouraged in Zambia and the regions, clinicians and policymakers should be well-versed in their strengths and limitations. We recommend a lower APRI threshold (0.5 or 0.65) for Zambia and consideration of the FIB-4 index at >3.25. Finally, alcohol screening and consideration should also be part of the HBV evaluation to determine when to start antiviral therapy in patients with chronic hepatitis B infection.

## Data Availability

Data from this study are a property of the government of Zambia and cannot be made publicly available. All interested readers can access the data set from the Chairperson of the Zambia Biomedical Research Ethics Committee or the National Health research Authority from the following contact details: The chairperson of the University of Zambia Biomedical Research Ethics Committee contact details: Ridgeway campus P.O Box 50110, Lusaka: Telephone:260-1-256067Fax:+260-1-250753 and the chairperson of the National Health research Authority contact details:Paediatric Centre of Excellence, University Teaching Hospital, P.O. Box 30075, LUSAKAChalala Office Lot No. 18961/M, Off Kasama Road, P.O. Box 30075, LUSAKA Tell: +260211 250309 | | www.nhra.org.zm

## Acknowledgements

Micah Benwa for helping in patient recruitment.

All the patients who agreed to participate in the study.

